# A Covid-19 case mortality rate without time delay systematics

**DOI:** 10.1101/2020.03.31.20049452

**Authors:** Richard Lieu, Siobhan Quenby, Ally Bi-Zhu Jiang

**Affiliations:** Department of Physics, University of Alabama, Huntsville, AL 35899, USA; Division of Reproductive Health, Warwick Medical School, The University of Warwick, UK; Shenzhen RAK wireless Technology Co., Ltd., China

## Abstract

Concerning the two approaches to the Covid-19 case mortality rate published in the literature, namely computing the ratio of (a) the daily number of deaths to a time delayed daily number of confirmed infections; and (b) the cumulative number of deaths to confirmed infections up to a certain time, both numbers having been acquired in the middle of an outbreak, it is shown that each suffers from systematic error of a different source. We further show that in the absence of detailed knowledge of the time delay distribution of (a), the true case mortality rate is obtained by pursuing method (b) at the end of the outbreak when the fate of every case has decisively been rendered. The approach is then employed to calculate the mean case mortality rate of 13 regions of China where every case has already been resolved. This leads to a mean rate of 0.527 ± 0.001 %.

---

In a recent correspondence to Lancet [1], the global case mortality rate of the coronavirus Covid-19 ([2]) was re-calculated after correcting for the finite time delay between diagnosis of the disease and death, which led to a higher estimate of the rate, namely 5.7 ± 0.2 % for the date of March 1, 2020, to be compared to the global mean value of 3.43 ± 0.01 % as computed from the data in [3, 4] for the same day. The reason for the higher value in [1] is the authors’ definition of the case mortality rate, as the ratio of the number of case-related deaths for the day of interest to the number of new confirmed infections for the same 1-day period two weeks earlier (obviously, this assumes distribution of time delay between infection and death is peaked at 14 days with a spread of less than 1 day). The usual definition, on the other hand, is the ratio of the cumulative number of deaths to confirmed infections, both being counted to the date of interest. If the daily number of confirmed infections and deaths are constants, the two definitions will give the same answer. This is no longer so if both numbers vary. Thus, if *e*.*g*. if they both increase with time but the former more steeply, the method of [1] will yield a larger result because the ratio involves a smaller denominator as a result of the smaller number of confirmed infections at an earlier time.

The purpose of this paper is to point out the respective limits of validity of the two approaches above, and under what circumstance would one be able to infer the true case mortality rate without assuming any details about what could happen after an infection is confirmed. We then apply our formalism to calculate the mean case mortality rate of several parts of China, which is completely free from the systematic error arising from the uncertain time delay between diagnosis and death. Our results for these regions do not corroborate the two high case mortality rates quoted above.

To begin with, we enlist the three quantities which are relevant to the calculation of the case mortality rate. First is the number of deaths per unit time *N* (*t*), second is the number of confirmed infections per unit time *n*(*t*), and third is the probability per unit delay time *p*(*t*) of a person dying at time *t after* she was diagnosed as a confirmed infection. More precisely *N* (*t*)*dt* and *n*(*t*)*dt* are respectively the number of deaths and the number of confirmed infections between the times *t* and *t* + *dt* from some arbitrary time origin before the outbreak of the disease, and *p*(*t*)*dt* is the probability of a person dying between the times *t* and *t* + *dt from* the time of diagnosis.

Evidently the three quantities *N, n*, and *p* must obey the relation

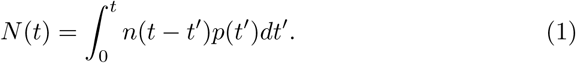

But since

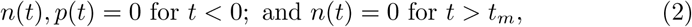

where *t*_*m*_ is the time beyond which no new infections are reported, it is possible to rewrite (1) as

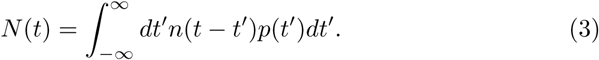

The case mortality rate is then given by

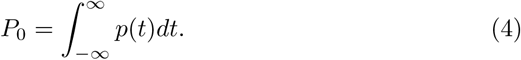

It can also be seen from (3) that *N* (*t*) is just the convolution integral between the two functions *n*(*t*) and *p*(*t*). We now explore two scenarios.

The first scenario is when the time delay between diagnosis and death has the unique value *t* = *t*_0_, so that *p*(*t*) = *p*_0_*δ*(*t − t*_0_) where *δ*(*t*) is the Dirac delta function and (from (4)) *p*_0_ = *P*_0_ is the case mortality rate being sought. In this limit, the integral (3) may readily be evaluated to yield

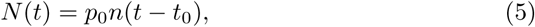

which means *p*_0_ is exactly the ratio defined by [1].

Under the second scenario, suppose one computes the cumulative number of deaths throughout the entire epidemic, as

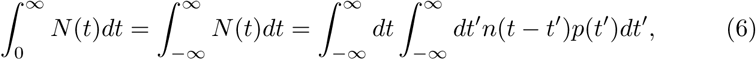

where use was made of (1), (2), and (3). One can readily change the variable of the *t* integration from *t* to *τ* = *t − t*^*′*^ to show that

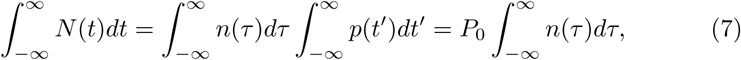

where use was made of (4).

Although (7) vindicates the conventional method of taking the ratio of the cumulative number of deaths to confirmed infections as the case mortality rate, beware that (7) applies *only* to the late time scenario when every case is recorded and its outcome (in terms of recovery versus death) is accounted for. In this respect, [1] is correct in asserting that the ratio does not yield the true probability of death if it is evaluated in the middle of an outbreak. Unfortunately, since the Covid-19 pandemic is far from over, and the assumption of a unique 14 day time delay between diagnosis and death is unrealistic, [5], neither [1] nor the conventional method would yield the true value of the case mortality rate.

**Table 1:**
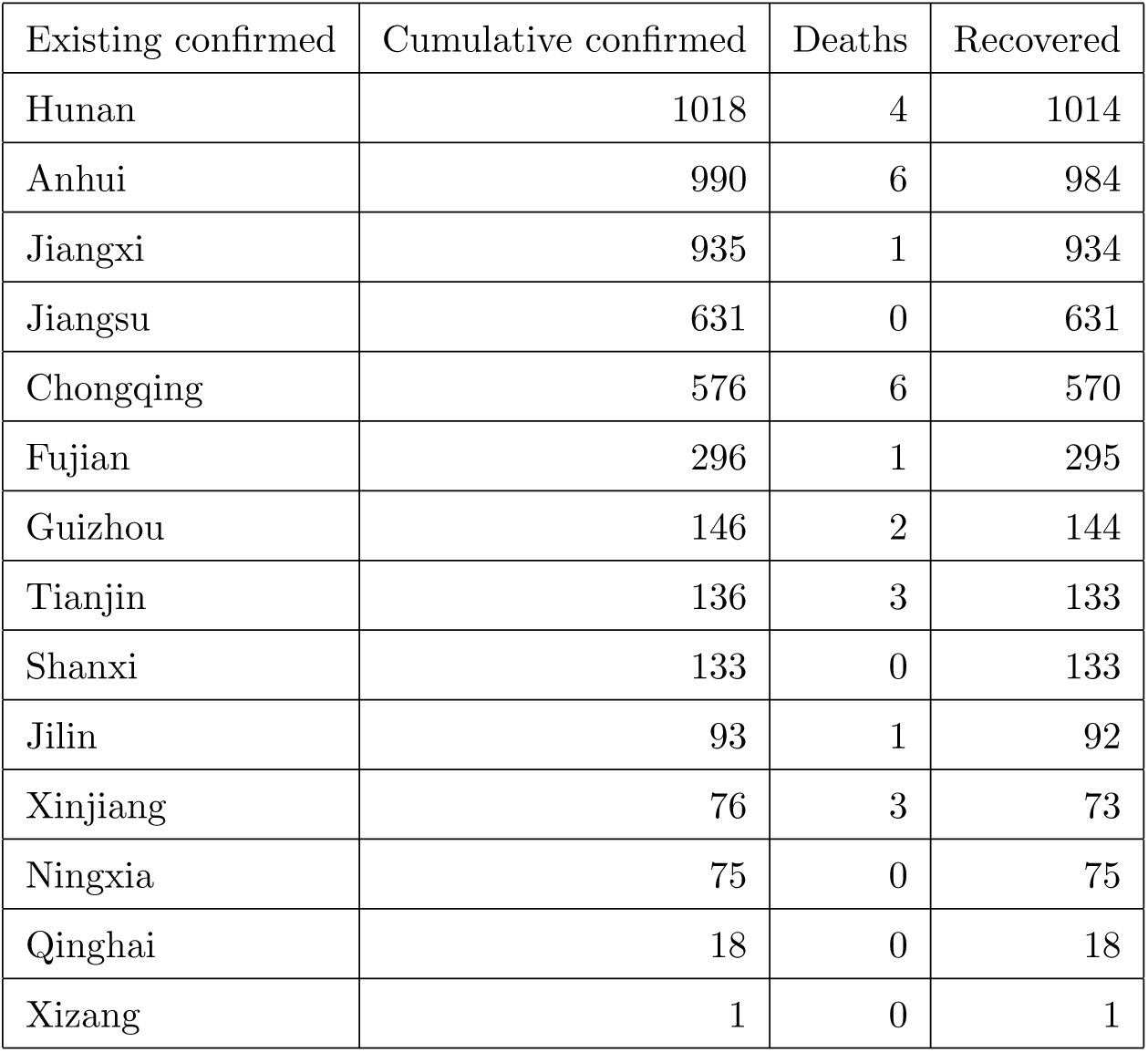
Number of Covid-19 related cases in each of 3 categories, and for 13 provinces of China[2, 6, 7] where every infected person has either recovered or died.

Nevertheless, even at the current stage of the Covid-19 outbreak there are regions of China in which the verdict of every confirmed case of infection has been delivered by the date of 16 March, 2020. The data for these regions (and their sources) are tabulated below.

Tallying the numbers, one obtains the case mortality rate *P*_0_ in accordance with (7) as *P*_0_ = 0.527 ± 0.001 %, where the error is due to random Poisson 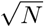 counting uncertainties in the cumulative death count *N* = 27. This is the true mortality rate for the regions of concern, which is free from the uncertainty in the time delay between diagnosis and death, but ignores the asymptomatic cases of infection. Evidently the rate is considerably lower than the two less accurate approaches quoted at the beginning of the paper, although the reason could be the smaller number of confirmed cases in these regions, which allows each patient to receive more attentive and higher quality health service.

## Data Availability

All the data used by this preprint are in the public domain (e.g. WHO, JHU, Worldometer).

